# HOW ARE RAPID RESPONSE SERVICES IMPLEMENTED OR CHANGED, AND HOW IS THEIR SUCCESS MEASURED?

**DOI:** 10.1101/2024.01.01.24300694

**Authors:** R. Rowley, A. L. Poulter, A. Smith, E. Pollock, D. Bush, P. Patel, M. Lam, L. Webb, D. Jones, A. Delaney

**Author notes:** Corresponding Author* Dr Rebecca Rowley.

## Abstract

**Introduction:** Rapid Response Teams (RRT) exist in many different formats. With escalating rapid response calls, RRT services need to adapt to meet demand. We will describe how RRT service changes are implemented, either as a novel service or service redesign, and how the success of the implementation is measured.

**Methods and analysis:** We will systematically review observational cohort studies that involve RRT implementation service change, and measure their implementation success. We will extract data relating to the study characteristics, team characteristics, methods of change implementation, and the outcome measures. The analysis will be primarily narrative, and we will present simple statistics regarding the range and frequency of the methods of implementation.

**Ethics and dissemination:** As this review is of published studies, it does not require ethical approval. We aim to present our results at scientific meetings and publish the manuscript to a peer reviewed journal.

**Trial registration number:** This protocol will be registered on the preprint server medRxiv.

**Strengths and limitations of this study:** The review addresses a relevant question, and will provide a comprehensive evaluation of the implementation of RRT services, and their measures of outcomes. It will serve to provide a basis for future study. We acknowledge that there will be limitations, including heterogeneity of eligible studies, such as variability in team name, composition, resource base, and differing outcome reporting. Additionally, it may be limited by the lack of studies relating to the implementation of services/change management.

## INTRODUCTION

### Rationale

A Rapid Response Team (RRT) is an acute hospital service which is activated when a patient is at risk of, or has deteriorated, requiring prompt medical intervention, with the goal of providing the right treatment at the right time for the right patient(1, 2). RRTs originated from Medical Emergency Teams (MET) concept, which were first described and introduced in 1990, at Liverpool Hospital, NSW, Australia. They arose due to the observation time leading up to cardiac arrest, and that recognising and intercepting deteriorating patients could improve outcomes. The goal of the team was to reduce in hospital cardiac arrests, hospital mortality, adverse events, and emergency ICU admission.

Nomenclature for this service is now variable, with RRT, MET, patient at risk team (PART) and Critical Care Outreach Team (CCOT) being the most common and often being used interchangeably(3). For the purpose of this protocol, we will use the term RRT to represent all related nomenclature.

Since their introduction, there has been many iterations of the RRT, dependent on (but not limited to):

- Size of hospital
- Services available at hospital
- The base service from which the service is branched from (ie ICU led, ED led, parent unit led)
- The funding allocated to the service.

The sparsest of services is led by a solitary nurse, whereas those with a dedicated department (for example, ‘Safety Afterhours for Everyone’ (SAFE) team in Perth, Western Australia) have a mixture of medical and nursing expertise. There are also Nurse Practitioner led services (Liverpool Hospital, NSW). The lead department is often variable, with ED and ICU most commonly taking ownership in Australia, however there are some services where the RRT is its own independently funded department.

The use of the service is similarly broad and undefined. What is unified across all variations is the need to attend to cardiac arrests, or acutely deteriorating patients; the defining features of “acute deterioration” is similarly varied, with over 30 different sets of calling criteria. Between The Flags (single calling criteria) and Early Warning Scores (combined observation parameters) are the most commonly used safety nets for capturing patients in clinical crisis(1, 3, 5, 10-13).

Given the complexity and heterogeniety of RRT research, it is not surprising that there is limited availability of a unifying structure regarding the best model, and limited resources and evidence for best adapting a service (for example in the context of funding modification, hospital redevelopment). With ongoing expansion of hospitals, increasing numbers of rapid response calls, and strain on RRT services, this lack of guidance and structural unity may prove challenging. Using this systematic review, we aim to explore how RRT services are implemented (defined as novel services in hospitals which did not previously have a RRT), or redesigned (defined as hospitals which previously had a service but have implemented resource or procedural change), why they are changed, over what time frame these change occur, and what parameters are used as measures of success. This will clarify what evidence is available, and highlight gaps for future study.

## METHODS

This systematic review will include prospective and retrospective observational cohort studies in line with the Cochrane Handbook of Systematic Reviews of Interventions recommendations, and will report our findings in accordance with the Preferred Reporting Items for Systematic Review and Meta-Analysis (PRISMA) statement(14). It has been registered on the preprint server MedRxiv.

### Eligibility Criteria

1. Study types: We will include prospective and retrospective observational cohort studies. There will be no restriction on publication status, language, nor publication year.
2. Population: Our population will be healthcare systems that are implementing a novel RRT service, or have an already established RRT service and are implementing service redesign.
3. Intervention: We will include studies where the intervention is the implementation or redesign (as defined above) of a RRT service.
4. Comparison: Inclusion will not be limited by a comparator.
5. Outcomes: We will include studies that report any measure of outcome.

### Exclusion Criteria

1. Studies that cannot be translated into English

### Information Sources

We will perform a search of the electronic databases: Medline (OVID), CINAHL (EBSCO), Embase (OVID), and The Cochrane Library (WILEY). Additionally, we will supplement our search with Google Scholar, with a limit of the first ten pages of results. We will manually search reference lists for eligible studies and other systematic reviews, and abstracts from relevant conferences, and contact experts in the field.

### Search Strategy

Our search strategy will be developed in keeping with the Peer Review of Electronic Search Strategies (PRESS) guideline statement(15). We will conduct MeSH and keyword searches for RRT services, service implementation and service redesign.

## STUDY RECORDS

### Data management

We will use COVIDENCE as our central store of study references. Data will then be extracted into a design specific excel spreadsheet.

### Selection process

Records identified will be downloaded into COVIDENCE, and duplications removed. Screening for potentially eligible studies will the take place by two independent authors, with any differences resolved by a third reviewer. Once screening is complete, we will retrieve all manuscripts identified as potentially eligible. The full texts will then be reviewed for eligibility by two authors independently and in duplicate. Reasons for exclusion will be recorded in a Preferred Reporting Items for Systematic Reviews and Met-analysis (PRISMA) flow diagram.

### Data collection process

The authors will develop a data extraction form, with a calibration exercise to test and refine the data collection form prior to formal collection. The authors will then independently and in duplicate extract data from each included study. Any unresolved queries will be rectified by seeking clarification from study authors. Any disagreement between reviewers will be resolved via discussion, then via a third reviewer.

### Data Items

We will extract data regarding:

1. Study characteristics
  ° first author
  ° year of publication
  ° study type
  ° number of participants
  ° location
  ° number of sites
  ° population
2. Team characteristics (pre + post if study reports service charge rather than novel)
  ° nomenclature of team
  ° constituents of team
  ° hours of service
  ° Calling system
  ° RRC/1000 separations
  ° IHCA/1000 separations
  ° early warning score (EWS) vs single parameter track and trigger or other
3. Implementation methods
  ° Novel service or redesign
  ° reason for change
  ° time frame of project implementation
  ° use of translational/ orientational simulation/technologies
  ° information dissemination techniques
4. System outcome assessments
  ° outcome measures of success
  ° method of ascertainment
  ° timing of follow-up
  ° duration of follow-up
  ° Quantative measures
  ° Qualitative measures

## OUTCOMES AND PRIORTISATION

### Primary

To describe the methods of implementation of RRT services, either as a novel service or as a service redesign.

### Secondary

1. To report why services are implemented or redesigned.
2. To report the time frame over which RRT services, or changes, are implemented.
3. To report key measures of outcomes for the implementation or redesign of MET services.

### Risk of bias in individual studies

We will use the JBI critical appraisal checklist for systematic reviews to assess risk of bias. It will be performed in duplicate with differences to be resolved by discussion.

### Data synthesis

Results will be mainly narrative, with any subsequently identified required analysis performed using the statistical software SPSS. We will present simple statistics regarding the range and frequency of the methods of implementation

## Data Availability

All data produced in this review are already publicly available, and will be available upon reasonable request to the authors

## ETHICS AND DISSEMINATION

As this review is of published studies, it does not require ethical approval. We aim to present our results at scientific meetings and publish the manuscript to a peer reviewed journal.

## DISCUSSION AND LIMITATIONS

This systematic review will provide a comprehensive review of the implementation of RRT services, and their measures of outcomes. It will serve to provide a basis for future study. We acknowledge that there will be limitations, including heterogeneity of eligible studies, such as variety in team name, constitution, resource base, and differing outcome reporting. Additionally, it may be limited by the lack of studies relating to the implementation of services/change management.

## FUNDING

There is no external funding for this review.

## Notes

### Competing Interest Statement

The authors have declared no competing interest.

### Funding Statement

This study will not receive any funding

### Author Declarations

The systematic review will include published studies only, therefore only using data that are openly available to the public.

